# Evoked midfrontal activity predicts cognitive dysfunction in Parkinson’s disease

**DOI:** 10.1101/2022.07.26.22278079

**Authors:** Arun Singh, Rachel C Cole, Arturo I Espinoza, Jan R. Wessel, James F. Cavanagh, Nandakumar S Narayanan

**Author notes:** **Corresponding Author** Nandakumar Narayanan, 169 Newton Road, Pappajohn Biomedical Discovery Building—5336, University of Iowa, Iowa City, 52242. **Co-Corresponding Author** Arun Singh, 414 E. Clark St., Sanford School of Medicine, University of South Dakota, Vermillion, 57069.

## Abstract

Cognitive dysfunction is a major feature of Parkinson’s disease (PD), but the pathophysiology remains unknown. One potential mechanism is abnormal low-frequency cortical rhythms which engage cognitive functions and are deficient in PD. We tested the hypothesis that midfrontal delta/theta rhythms predict cognitive dysfunction in PD. We recruited 100 PD patients and 49 demographically-similar control participants who completed a series of cognitive control tasks, including the Simon, oddball, and interval timing tasks. We focused on cue-evoked delta (1-4 Hz) and theta (4-7 Hz) rhythms from a single midfrontal EEG electrode (Cz) in PD patients who were either cognitively normal, with mild-cognitive impairments (PDMCI), or had dementia (PDD). We found that PD-related cognitive dysfunction was associated with increased response latencies and decreased midfrontal delta power across all tasks. Within PD patients, the first principal component of evoked EEG features from a single electrode (Cz) strongly correlated with clinical metrics such as the Montreal Cognitive Assessment (MOCA; rho=0.36) and with NIH-toolbox Executive Function scores (rho=0.46). These data demonstrate that cue-evoked midfrontal delta/theta rhythms directly relate to cognition in PD. Our results provide insight into the nature of low-frequency frontal rhythms and suggest that PD-related cognitive dysfunction results from decreased delta/theta activity. These findings could facilitate the development of new biomarkers and targeted therapies for cognitive symptoms of PD.

## Introduction

Parkinson’s disease (PD) is a neurodegenerative disorder that features prominent cognitive symptoms, affecting up to 80% of patients at some point in the disease.^1,2^ PD degrades cognitive function^3,4^ leading to mild cognitive impairment (PDMCI) or dementia in PD (PDD).^5^ There are few reliable treatments for these aspects of PD because the mechanisms are not well understood.

Cognitive dysfunction in PD can impair executive processes such as conflict resolution, attention, and timing.^6–8^ Direct neurophysiological measurement via intracranial recordings or scalp electroencephalography (EEG) has identified prefrontal neural systems underlying these cognitive functions,^9–12^ whose activity is particularly signified by low-frequency oscillations in the delta (1-4 Hz) and theta (4-7 Hz) bands located over midfrontal regions.^13^ Midfrontal delta/theta rhythms are reliably modulated by conflict, attention, and timing,^13,14^ and specifically impaired in PD.^14–18^ Critically, midfrontal low-frequency rhythms engage single neurons in distributed cortical and subcortical brain networks that are involved in specific cognitive operations, such as conflict resolution, response inhibition, timing, and error correction.^18–23^ This line of work motivates the hypothesis that deficits in midfrontal delta/theta rhythms underlie cognitive dysfunction in PD.

We tested this idea within a large sample of PD patients across a range of cognitive functions, including mild-cognitive impairment (PDMCI) and dementia (PDD), as well as within demographically-equivalent controls. We used three well-described cognitive-control tasks that are reliably associated with impairments in midfrontal delta/theta rhythms in PD: 1) the Simon-conflict task,^17,24–27^ 2) an oddball task,^16,18^ and 3) an interval-timing task.^14^ These tasks reliably trigger low-frequency midfrontal EEG rhythms that can be captured at a single electrode located at the cranial vertex (Cz).^13,14,16–18,21,28,29^ Midfrontal delta/theta rhythms are proposed to signal a need for cognitive control^12,29,30,30,31^ during simple responses to novelty^18^ and executive functions.^14,32^ We report three main results. First, across all three tasks response time was progressively affected by PD-related cognitive function (PD<PDMCI<PDD). Second, midfrontal delta rhythms were progressively attenuated by PD-related cognitive dysfunction. Finally, principal component analyses identified common midfrontal delta/theta variance across all tasks, and this single factor strongly correlated with clinical cognitive metrics and could be used to classify PDD. These data identify frontal mechanisms of cognitive dysfunction in PD and could inform new biomarkers and treatments for neurodegenerative disease.

## Materials and methods

### Participants

We recruited a total of 100 PD patients and 49 controls to perform a variety of cognitive and motor tasks from the University of Iowa Movement Disorders Clinic between 2017 and 2021.

All patients were examined by a movement-disorders physician to verify that they met the diagnostic criteria recommended by the United Kingdom PD Society Brain Bank criteria. Control participants were recruited from the Iowa City community and similar in terms of age, sex, and education. All PD and control participants were determined to have the decisional capacity to provide informed consent in accordance with the Declaration of Helsinki and the Ethics Committee on Human Research. We obtained written informed consent from each participant. All research protocols were approved by the University of Iowa Human Subjects Review Board (IRB# 201707828). Data from some of these patients have been published previously,^14,18^ but we did not evaluate cognitive function across tasks in detail, which is our focus here. All data were collected with patients on levodopa as usual, as previous work has reliably demonstrated that levodopa does not reliably influence midfrontal low-frequency rhythms.^14,17,33,34^ We used the Montreal Cognitive Assessment (MOCA) to stratify cognitive function in PD.^5,35^ We defined (PDD) dementia as MOCA <22; mild-cognitive impairment (PDMCI) as MOCA 22-26; and cognitively normal as MOCA 26-30 (referred to as PD or control). All data were collected during a single 4–5-hour session with EEG. Patients also performed the full UPDRS and Executive Function tests from NIH Toolbox Cognition Battery, which included the Eriksen Flanker Test and the Dimension Change Card Sort.^36,37^ Patients were excluded for electrical noise, if they could not perform enough trials, technical issues, or fatigue. Demographics of recruited patients and control subjects are summarized in Table 1.

**Table 1:** Demographic, disease, non-motor, motor, and cognitive characteristics.

|  | Control (N=49) | PD (N=47) | PDMCI (N=34) | PDD (N=19) |
| --- | --- | --- | --- | --- |
| **Demographics and Disease** |  |  |  |  |
| Gender, M/F | 26/23 | 27/20 | 25/9 | 16/3 |
| Age, years | 70.9 (1.1) | 66.0 (1.1) | 71.2 (1.4) | 70.2 (1.9) |
| Disease duration, years | - | 4.8 (0.6) | 4.4 (0.5) | 4.8 (1.0) |
| LEDD, mg/day | - | 758.8 (54.9) | 750.3 (75.3) | 1065.8 (130.5) |
| **Cognition Characteristics** |  |  |  |  |
| MOCA (0-30) | 26.7 (0.3) | 27.4 (0.2) | 23.7 (0.2) | 17.8 (0.8) |
| **Motor Characteristics** |  |  |  |  |
| UPDRS III (0-56) | - | 11.2 (1.0) | 11.9 (1.1) | 16.5 (1.9) |

Values are expressed as mean (standard error of the mean).
Abbreviations: Male, M; Female, F; Levodopa Equivalent Daily Dose, LEDD; Montreal Cognitive Assessment, MOCA; motor Unified Parkinson's Disease Rating Scale, UPDRS III.
<sup>a</sup>Mann-Whitney U test was used for comparison between PD vs PDDEP subjects. Abbreviations: Male, M; Female, F; Montreal Cognitive Assessment, MOCA; Geriatric Depression Scale, GDS; Patient Health Questionnaire-9, PHQ-9; motor Unified Parkinson's Disease Rating Scale, UPDRS III.

#### Apparatus

All participants sat in a quiet room in front of a computer for behavioral assays during EEG recordings. All task stimuli were presented via the PsychToolbox-3 functions^38^,39, http://psychtoolbox.org/,^40^ in MATLAB on a Dell XPS workstation with a 19-inch monitor. Task-specific audio was played through Dell Rev A01 speakers positioned on either side of the monitor, and responses were made with the left and right index fingers on a standard QWERTY USB-keyboard. They performed the Simon task, oddball task, and interval timing task. Each of these paradigms have been extensively studied from the perspective of cognitive control and PD, and were specifically selected to study cognitive control in PD.^15,16,18,41–43^

#### Simon task

This and all subsequent tasks were presented using Psychtoolbox-3. 48 control, 47 PD, 34 PDMCI, and 18 PDD subjects were able to complete the Simon task (Fig. 1A). Briefly, each stimulus was presented to the left or right side of the screen, and participants were instructed to press a left key when the stimulus was yellow or red and a right key when it was cyan or blue. These presentations were thus either spatially congruent with the screen side matching the response hand (i.e., right-sided stimulus and responding with the right hand), or incongruent, with the screen side contralateral to the response hand (i.e., right-sided stimulus and responding with the left hand) as in a standard Simon task. Stimuli consisted of four randomly assigned unique shapes. There were 120 congruent and 120 incongruent trials presented randomly in 4 blocks of 60. Data from congruent and incongruent trials were analyzed separately.

**Figure 1.**
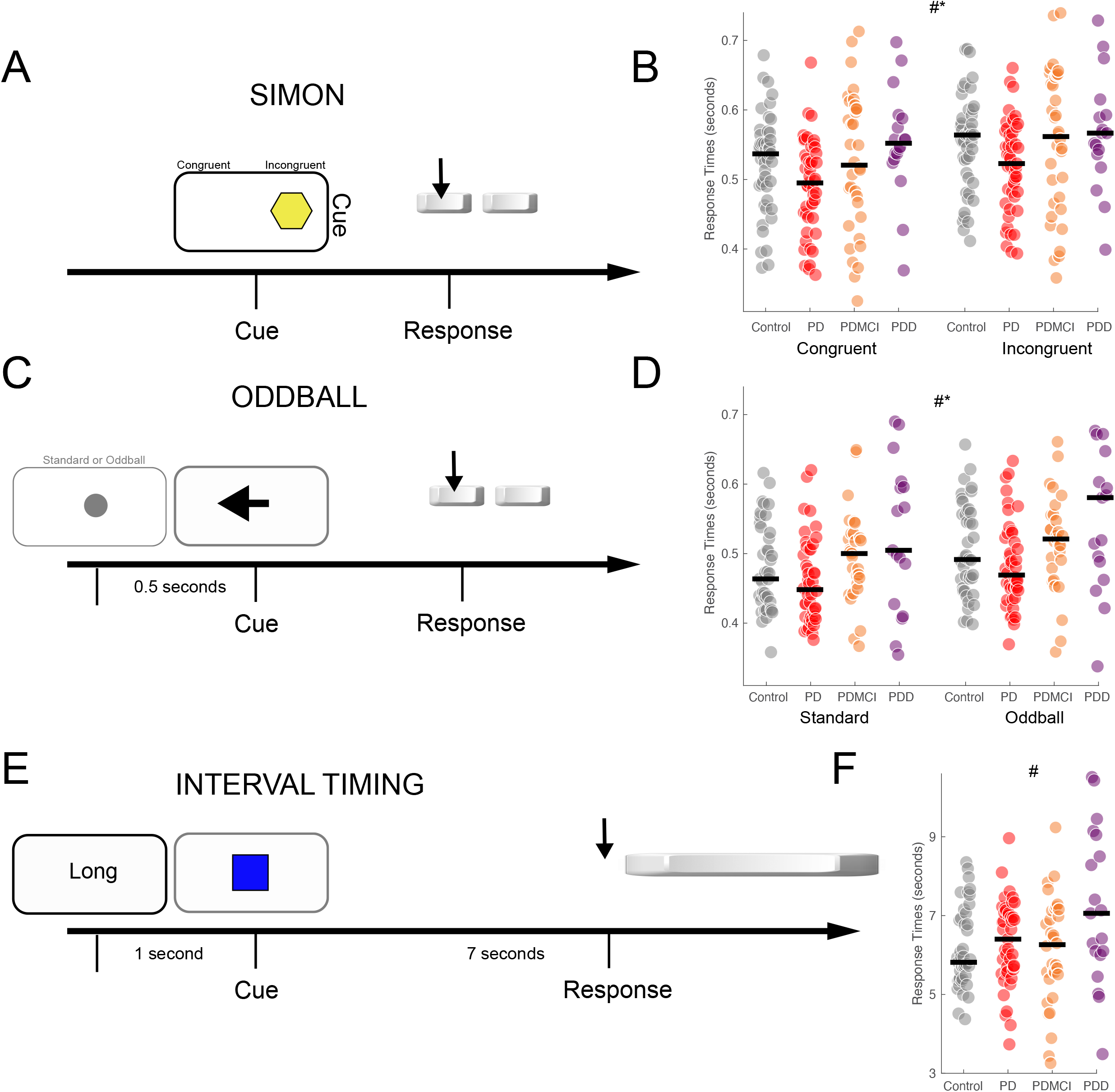
Responses during cognitive control tasks are slowed with cognition in PD. A) Simon task, in which patients are presented cues on the left or the right of the screen which indicates a left or right response. Congruent trials involve a cue on the same side (i.e., left cue on the left) and incongruent trials involve cues on opposite side (left cue on the right). B) Response times for congruent and incongruent trials for 48 control (gray), 47 PD (red), 34 PDMCI (orange), and 18 PDD (plum) patients; the black bar is the median. C) Oddball task, in which standard stimuli (80% of trials) and novel auditory cues (10% of trials) precede a left or right response as cued by an arrow. 10% of trials included a novel visual cue; these were not analyzed. D) Response times for standard and oddball trials for 41 control, 43 PD, 29 PDMCI, and 18 PDD patients; the black bar is the median. E) Interval timing task in which participants must estimate an interval of 7 seconds with a keypress. F) Response times for 43 control, 43 PD, 32 PDMCI, and 19 PDD patients; the black bar is the median. *=main effect of trial Type; #=main effect of Group.

#### Oddball task

We modified a cross-modal oddball distractor task, as described in detail previously.^18,44–46^ 41 control, 44 PD, 29 PDMCI, and 17 PDD subjects were able to complete the oddball task. A white arrow appeared in the center of a black screen, and the participant was required to press the key that corresponded with the direction of the arrow (“q” for left arrow, “p” for right arrow) as quickly as possible. The appearance of the arrow was preceded by an audio-visual cue by 0.5 seconds (either the standard cue or the distractor cue). Participants were instructed that this cue would appear 0.5 seconds before the target stimulus (white arrow), and that this cue would be a green circle and a short tone (600-Hz sine wave tone lasting 0.2 seconds). The audio-visual cue was followed by the target arrow (Fig. 1B). Participants had to respond within 1 s, after which the fixation cross reappeared and the next trial started after a variable inter-trial interval between 0.5 – 1 second. Participants completed a brief practice of the task (10 trials), which had all the familiar standard cues described above. Participants completed four blocks of 60 trials each, for a total of 240 trials. Of the 240 trials, 80% contained the standard cues as described above, 10% contained an unexpected auditory oddball cue (non-repeating, randomly-created sine wave that sounded like a birdcall or robotic noise lasting 0.2 seconds in duration) in place of the expected tone, and 10% contained an unexpected visual oddball cue which were not analyzed. Data from standard and oddball trials were analyzed separately.

#### Interval-timing task

We performed a modified version of the peak-interval-timing task as described in detail previously.^14,41^ 43 control, 43 PD, 32 PDMCI, and 19 PDD participants estimated the duration of an interval after reading instructions displayed as text at the center of a video screen. White instructional text on the center of a black screen was presented that read “Short interval” on 3-second interval trials and “Long interval” on 7-second interval trials. The actual interval durations were never communicated to the patient; we only analyzed long interval trials in this manuscript because our previous study observed no significant differences between PD and control subjects during short interval trials.^14,41^ After the instructions were displayed for 1 second, the start of the interval was indicated by the appearance of an image of a solid box in the center of the computer screen (Fig. 1C), which was displayed for the entire 18–20 seconds for 7-second intervals. Participants were instructed to press the keyboard spacebar at the start of when they judged the target interval to have elapsed (Fig. 1C). Participants were instructed not to count, and a distractor vowel appeared at random intervals in the screen center. All participants performed four blocks of 40 trials of randomly intermixed short and long intervals. Participants took a self-paced break between each block and moved to the next block by pressing any key.

### EEG Recordings

EEG recordings were performed according to methods described in detail previously.^42,47^ Briefly, we used a 64-channel EEG actiCAP (Brain Products GmbH) with a 0.1-Hz high-pass filter and a sampling frequency of 500 Hz. We used electrode Pz as an online reference and electrode site Fpz for the ground. EEG activity was referenced according to the procedures described in Singh et al.^42,47^ An additional channel was recorded at the mid-inion region (Iz), and we removed unreliable FT9, FT10, TP9, and TP10 channels, resulting in 59 channels for pre- and post-processing. Data were epoched around the cues from -1 to 2.5 seconds peri-cue.

Bad channels and bad epochs were identified using a combination of the Fully Automated Statistical Thresholding for EEG artifact Rejection (FASTER) algorithm that rejects artifacts with greater than +/- 3 Z-scores on key metrics^48^ and the pop_rejchan function from EEGLAB. Bad channels were subsequently interpolated, except the midfrontal “Cz” channel was never interpolated. Eye blink contaminants were removed following independent component analysis (ICA).

Event-related potentials (ERPs) were low-pass-filtered at 20 Hz for analyses. We calculated average ERP for each trial type and each group (PD and control). ERPs were visually inspected, and we calculated the mean amplitude for largest deflections at electrode Cz. This included the P3 component for the Simon (0.4-0.5 seconds from cue) and oddball task (0.25-0.35 seconds), as well as the N1 for the oddball task (0.05-0.15 seconds), and the first negative wave (0.15 – 0.35 seconds) for interval timing.

Our primary interest was in midfrontal delta/theta rhythms; consequently, we utilized time-frequency analyses. We computed spectral measures by multiplying the fast Fourier transformed (FFT) power spectrum of single-trial EEG data with the FFT power spectrum of a set of complex Morlet wavelets (defined as a Gaussian-windowed complex sine wave: e^i2πtf^e^-t^2/(2xσ^2)^, where *t*=time and *f*=frequency). Wavelets increased from 1–50 Hz in 50 logarithmically-spaced steps, which defined the width of each frequency band, increasing from 3–10 cycles between 1–50 Hz and taking the inverse FFT.^49^ The end result of this process was identical to time-domain signal convolution, and it resulted in estimates of instantaneous power (the squared magnitude of the analytic signal) and phase angle (the arctangent of the analytic signal). We then cut the length of each signal accordingly for each trial (−0.5 – 1 seconds). These short temporal epochs reflect the wavelet-weighted influence of longer time and frequency periods. Power was normalized by converting to a decibel (dB) scale (10*log10(power_t_/power_baseline_)), allowing us to directly compare the effects across frequency bands. The baseline for each frequency was calculated by averaging power from -0.3 to -0.2 seconds prior to cue onset.^42,43,47^

We defined delta power as 1-4 Hz and theta power as 4-7 Hz for our time-frequency regions of interest (tf-ROI). These frequency bands were extremely well-justified based on extensive prior work from our groups.^13,18,21,29,42,43^ Temporal epochs were defined based on peaks in average spectral activity across all groups.^49^ For the Simon task, these epochs spanned 0.4-0.7 seconds for delta/theta bands; for the Oddball task, these spanned 0.6-0.9 seconds for delta and 0.1 – 0.2 seconds for theta; for the Interval timing task, these spanned 0.15 – 0.45 seconds for delta and 0.1-0.3 seconds for theta. Our primarily analyses focused on how activity in these tf-ROIs changed across PD patients with cognitive dysfunction.

### Statistical Analyses

For response times, event-related potentials (ERP), and mid-frontal delta/theta power, we used a linear model (*lm* in R) with main effects of Group (Control/PD/PDMCI/PDD) and trial Type (Congruent vs. Incongruent, or Standard vs. Oddball) and Group*Trial type interaction. Interval timing analyses only had Group-level analyses. Post-hoc comparisons were performed via estimated marginal means (*emmeans* in R) and Tukey’s post-hoc correction. Effect size was computed via eta^2^ *(etaSquared* in R).^50^

The correlations between response time, ERP components, and mid-frontal delta and theta power tf-ROIs were first investigated via Spearman’s rho. We then used principal component analyses (PCA) to distill the shared variance across these metrics. We correlated the first principal component and clinical variables such as MOCA and Executive Function scores from the NIH-Toolbox.^37^ Correlation coefficients were comparing using the *cocor* toolbox in R using Hittner’s modification of Fisher’s R to Z.^51^

Pattern classification was performed using support vector machines (SVM; *fitcsvm*.*m* in MATLAB) with 500 iterations of 5X, 10X, and leave-one-out (LOO) cross-validation. Feature selection leveraged the first 3 principal components (exploratory analysis suggested more components did not add more information). Predictions were computed based on an SVM model using *predict*.*m* in MATLAB. ROC curves were computed using *perfcurve*.m. Classification was done for 1) PD (n=40) vs. PDD (n=16), 2) PD vs PDMCI (n=43), and 3) PDMCI vs. PDD, and 4) between group shuffled data derived from the PD vs. PDD group. Statistical significance was assessed using an empirically-derived p-value obtained by 1000 repetitions of classification of group-shuffled data.

## Results

We investigated how cognition in PD affected cognitive control. First, we examined response times (RT) during the Simon, oddball, and interval timing tasks (Fig. 1). For the Simon task, there was a main effect of Group (Control, PD, PDMCI, and PDD; F=6.6, p=0.002, eta^2^=0.06; see table 2 for RTs for all tasks) as well as trial Type (congruent vs. incongruent; F=5.6, p=0.02, eta^2^=0.03), but no interaction (F=0.0, p=0.99; post-hoc comparisons were not significant). Accordingly, there was no main effect of Group on response slowing on incongruent vs. congruent trials (Supplementary Fig. 1). This pattern was similar in the oddball task, with a main effect of Group (F=9.3, p=0.0001, eta^2^=0.09) and a marginally significant effect of trial Type (oddball vs. standard; F=3.2, p=0.07, eta^2^=0.02). As in the Simon task, there was no interaction (F=0.1, p=0.92; post-hoc comparisons for PD vs. PDD were marginally significant for oddball p=0.07 and significant for standard p=0.04) and no main effect of Group in oddball-related slowing (F=0.3, p=0.78). These findings indicate that PD-related cognitive dysfunction does not reliably affect behavioral indicators of control adaptation during incongruent Simon trials or novelty during Oddball trials. Finally, there was a significant effect of Group in the interval timing task (F=4.2, p=0.02, eta^2^=0.09; post-hoc comparisons were significant between PD and PDD p=0.05 and between PDMCI and PDD p=0.02).

**Table 2:** Response times across tasks (in milliseconds)

| **SIMON** |  |  |  |  |  |  |
| --- | --- | --- | --- | --- | --- | --- |
|  | <u>Congruent</u> |  |  | <u>Incongruent</u> |  |  |
|  | *Median* | *Q1* | *Q3* | *Median* | *Q1* | *Q3* |
| Control | 537 | 490 | 561 | 558 | 509 | 592 |
| PD | 499 | 451 | 536 | 522 | 479 | 567 |
| PDMCI | 520 | 450 | 602 | 562 | 473 | 641 |
| PDD | 540 | 522 | 586 | 561 | 542 | 611 |

| **ODDBALL** |  |  |  |  |  |  |
| --- | --- | --- | --- | --- | --- | --- |
|  | <u>Standard</u> |  |  | <u>Oddball</u> |  |  |
|  | *Median* | *Q1* | *Q3* | *Median* | *Q1* | *Q3* |
| Control | 464 | 432 | 530 | 487 | 451 | 560 |
| PD | 445 | 412 | 505 | 469 | 435 | 525 |
| PDMCI | 498 | 451 | 528 | 522 | 484 | 553 |
| PDD | 506 | 426 | 596 | 533 | 461 | 644 |

| **INTERVAL TIMING** |  |  |  |
| --- | --- | --- | --- |
|  | *Median* | *Q1* | *Q3* |
| Control | 5889 | 5476 | 6918 |
| PD | 6437 | 5597 | 7054 |
| PDMCI | 6269 | 5455 | 7140 |
| PDD | 7118 | 6011 | 8911 |

We next turned to EEG, which captures cortical neurophysiology with fine temporal resolution, to explore the neural bases of these PD-related changes to behavior. Selected ERP components from the midfrontal electrode Cz did not reveal main effects of Group in Simon or timing tasks (Supplementary Fig. 2). However, for oddball-related activity between 0.25-0.35 seconds (Supplementary Fig. 2D-F), we found main effects of Group (F=5.6, p=0.005, eta^2^=0.04) as well as Type (oddball vs. standard; F=36.2, p=1×10^−8^, eta^2^=0.16), and a marginally significant interaction (F=2.6, p=0.08; post-hoc comparisons between PD and PDD for oddball p=0.002). This was also true for the earlier N1 peak (0.05 – 0.15 seconds; Supplementary Fig. 3): Group (F=4.4, p=0.01, eta^2^=0.05) and Type (F=4.3, p=0.04, eta^2^=0.02), but without an interaction. These data suggest that oddball-related ERPs could independently discriminate cognitive dysfunction in PD.^16,28^

**Figure 2.**
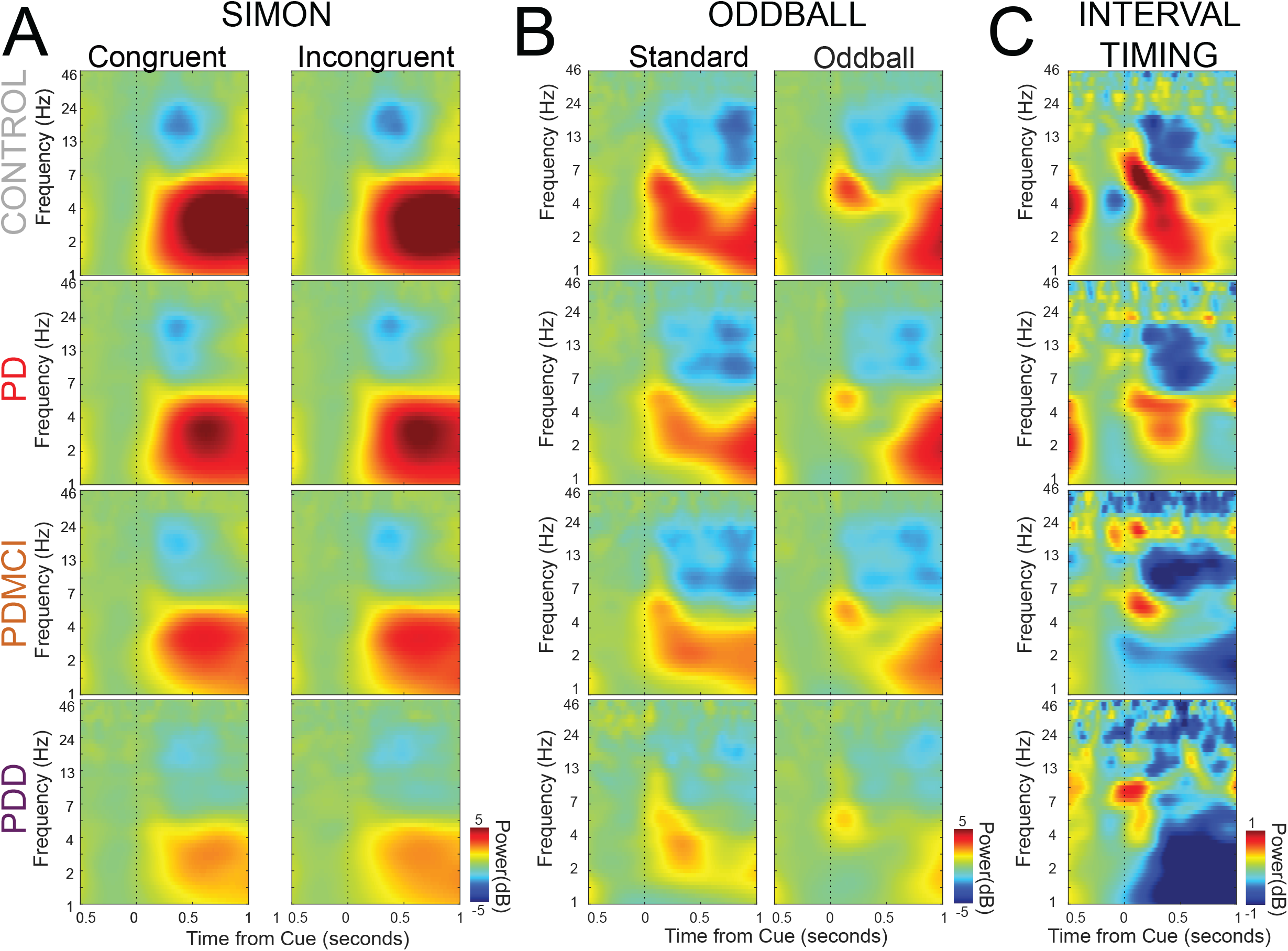
Midfrontal delta/theta rhythms in Simon, Oddball, and Interval timing tasks: A) Cue-evoked time-frequency spectrograms from EEG electrode Cz during the Simon task on congruent and incongruent trials for 48 control, 47 PD, 34 PDMCI, and 18 PDD patients. B) Cue-evoked time-frequency spectrograms from EEG electrode Cz during the Oddball task on Standard and Oddball trials for 41 control, 43 PD, 29 PDMCI, and 18 PDD patients. C) Cue-evoked time-frequency spectrograms from EEG electrode Cz during the Interval Timing task for 43 control, 43 PD, 32 PDMCI, and 19 PDD patients. Across tasks, there was a consistent pattern of decreased midfrontal cue-evoked delta/theta power.

**Figure 3.**
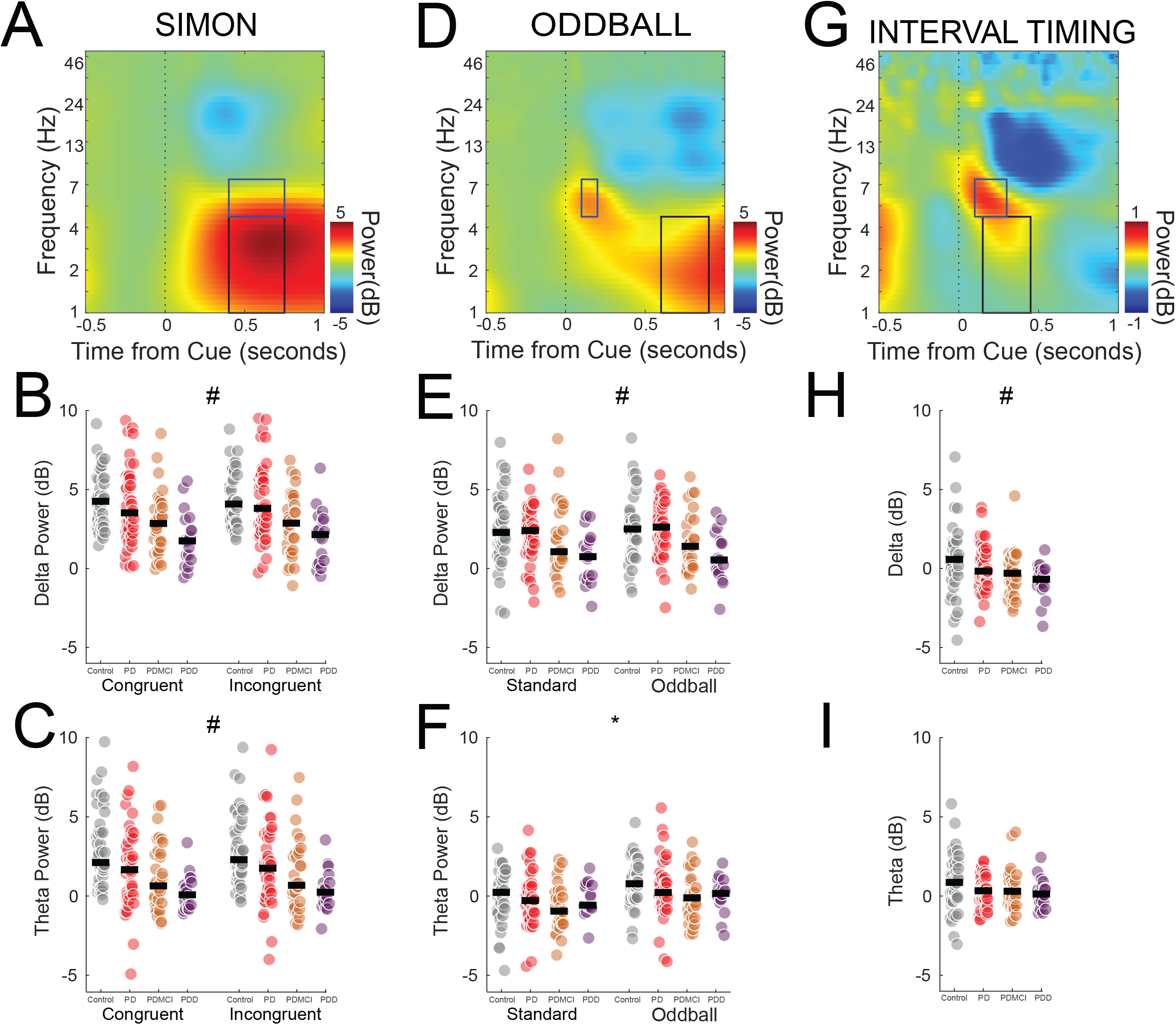
Cue-triggered midfrontal delta/theta activity is decreased with cognitive dysfunction in PD. We performed time-frequency analyses of activity from midfrontal electrode Cz. During the A) Simon task across all participants, we found prominent cue-triggered low-frequency rhythms <7 Hz. We identified time-frequency regions of interest (tf-ROIs; outlined in black) for theta (4-7 Hz) and delta (1-4 Hz) bands from 0.4– 0.7 seconds; 48 control (grey), 47 PD (red), 34 PDMCI (orange), and 18 PDD patients (plum). Power for delta (B) and theta (C) bands was reliably attenuated with cognitive impairments in PD. D) Oddball task time-frequency analyses revealed that E) delta bands from 0.6-0.9 seconds and F) theta bands from 0.1-0.2 seconds were impaired with cognition in PD. 41 control, 43 PD, 31 PDMCI, and 18 PDD patients. G) Interval-timing time-frequency analyses revealed that (H) theta bands from 0.1-0.3 seconds and I) delta bands from 0.1-0.45 seconds were also reliably impaired with cognition in PD; 43 control, 43 PD, 32 PDMCI, and 19 PDD patients; the black bar is the median. *=effect of trial Type; #=main effect of Group.

Our past work has suggested that spectral analyses of midfrontal EEG can predict cognitive dysfunction in PD.^15,42,43^ We tested this idea by applying time-frequency analyses to midfrontal activity during the Simon, oddball, and interval timing tasks. We focused on midfrontal EEG electrode Cz which past work has established as the focus of cognitive control.^18,21,29,42,43^ We noticed that across tasks, there was a consistent pattern of decreased cue-evoked midfrontal delta/theta power that decreased with cognitive dysfunction in PD (Fig. 2).

To quantify this finding, we selected midfrontal tf-ROIs. We focused on delta (1–4 Hz) and theta (4-7 Hz) bands that are extraordinarily well-justified from our prior work.^13,18,21,29,42,43^ Temporal epochs for tf-ROIs were based on maximal spectral activity on average across all patients (Fig 3A,3D, and 3G). Here we focused on how activity in these EEG tf-ROIs covaried with cognitive dysfunction in PD. For the Simon task, there were main effects of Group on delta power (1-4 Hz; 0.4-0.7 seconds; F=13.6, p=3×10^−6^, eta^2^=0.12; post-hoc comparisons between PD vs. PDD were significant for congruent p=0.01 and incongruent p=0.005) and theta power (4-7 Hz; 0.4-0.7 seconds; F=6.2, p=0.003, eta^2^=0.06), but no effects of trial Type or higher interactions (delta: F=0.0; p=0.98; theta: F=0.0, p=1.0). We observed a similar pattern for the oddball (Fig. 3D) delta ROI with main effects for Group (1-4 Hz; 0.6-0.9 seconds; F_=_8.5, p=0.003, eta^2^=0.09; post-hoc comparisons for PD vs. PDD for incongruent p=0.02; Fig. 3E) but no effect of Type or higher interactions (delta: F=0.1, p=0.88). However, for theta ROIs (4-7 Hz; 0.1-0.2 seconds; Fig. 3F), there was a main effect of Type (F=6.2, p=0.01, eta^2^=0.03) but were no main effects for Group (F=1.6, p=0.21) and no higher interactions (F=0.3, p=0.74). Finally, for Interval Timing (Fig. 3G), there were main effects of Group on delta power (Fig. 3H; 1-4 Hz; 0.15 – 0.45 seconds; F=4.2, p=0.02, eta^2^=0.09; post-hoc comparisons between PD vs PDD were significant p=0.01) but no reliable effects on theta power (Fig. 3I; 5-7 Hz, 0.1-0.3 seconds; F=0.2, p=0.83). Across tasks, these data provide convergent evidence that PD-related cognitive disturbance is consistently associated with diminished delta–band power (eta^2^=0.09-0.12). Surprisingly, with the exception of oddball-related theta power, there was no reliable evidence of control-related effects for congruent vs. incongruent or standard vs. oddball contrasts (Supplementary Fig. 4). Taken together, these data suggest that PD patients with cognitive dysfunction fail to engage cue-evoked delta band power.

**Figure 4.**
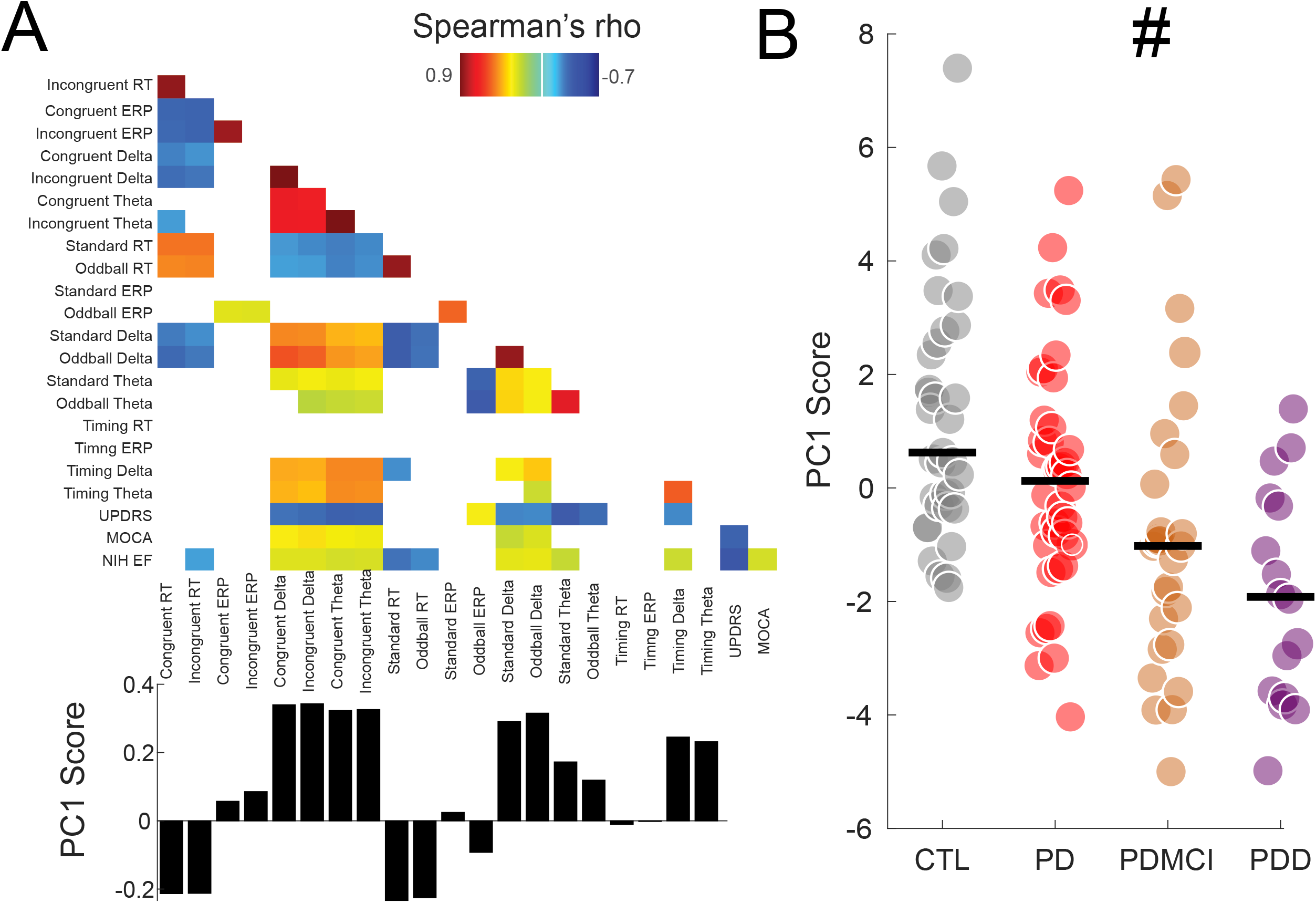
Principal components identify shared variance across tasks. Data from 34 control participants (grey), 40 PD (red), 27 PDMCI (orange), and 16 PDD participants (plum) who completed all tasks. A) Correlation matrix of response-times and delta/theta activity from Simon, Oddball, and interval timing tasks. Below, the first component from principal component analysis (PC1) loaded highly on delta/theta activity, and inversely on response time. B) PC1 was strongly affected by cognition in PD and PC1 explained 30% of variance in A. #=main effect of Group.

We examined the relationship between these metrics across tasks and patients. There were 35 control participants, 40 PD patients, 27 PDMCI patients, and 17 PDD patients that completed all tasks. Correlation values between these variables are shown in Fig. 4A, only including values with a corrected FDR p<0.05. We used a data-driven principal component analysis to capture patterns across task.^52^ PC1 loaded highly on delta and theta activity across tasks (Fig. 4B). Strikingly, we found that PC1 effectively captured differences in cognition in PD (Main effect of Group on PC1 Score: (F=5.1, p=0.008, eta^2^=0.11; post-hoc differences between PD and PDD p=0.008; Fig. 4B) and explained 30% of variance in our data across all tasks.

Shared midfrontal variance as captured by PC1 was strongly linked with clinical metrics of cognitive function in PD patients. Specifically, PC1 was correlated with cognitive function as measured by the MOCA (Spearman rho=0.34, p=0.002; Fig. 5A) and Executive Function from the NIH Toolbox (Spearman rho=0.46, p=0.00003; Fig. 5B). Of note, there was a significant negative correlation between PC1 and the UPDRS (Spearman rho=-0.23, p=0.03), suggesting that better motor function was linked with higher PC1 scores. The correlation between UPDRS and PC1 was significantly less than the correlation between EF and PC1 (z = 3.77, p-value = 0.0002). No significant correlations were found between PC1 score and MOCA and Flanker activity for controls (Supplementary Fig. 5).

**Figure 5.**
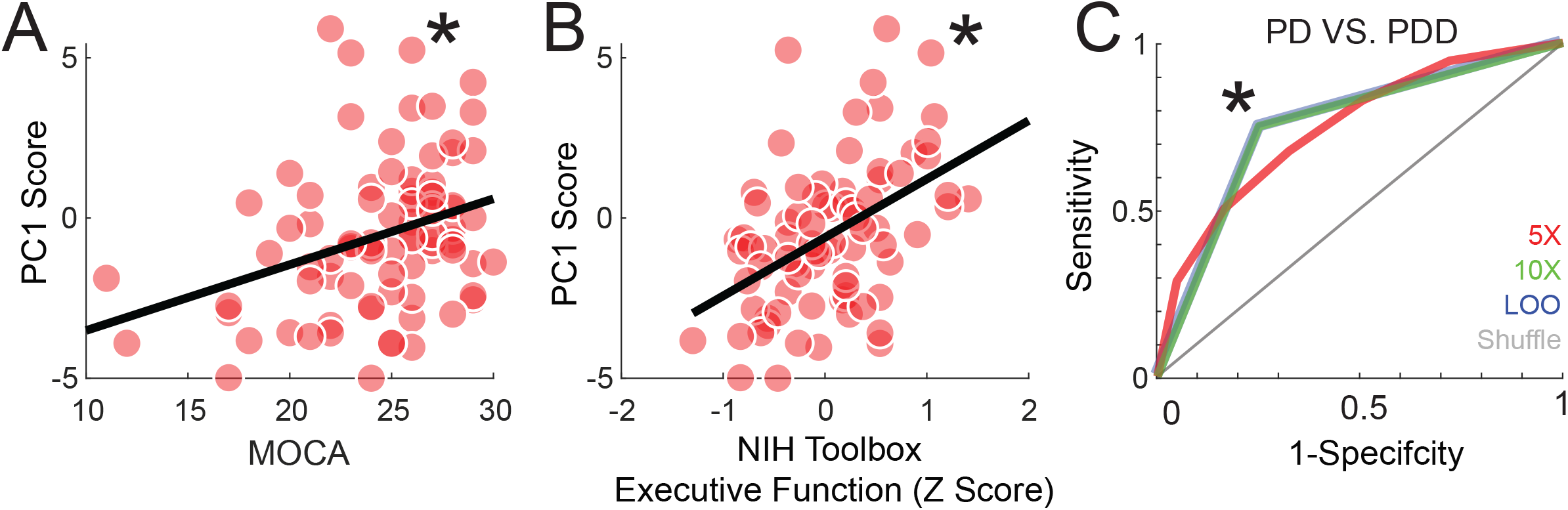
Shared midfrontal variance predicts cognitive dysfunction in PD. A) PC1 correlated with cognitive function, as identified by the Montreal Cognitive Assessment (rho=0.35) and by B) NIH Toolbox executive dysfunction (rho=0.46). No reliable correlation was observed in control participants. *=p<0.05. C) Pattern classification techniques could accurately identify PD patients without cognitive dysfunction (PD; 40 participants), or PD patients with cognitive dysfunction (PDD; 16 participants). 5x cross-validation in red, 10x cross-validation in green, leave-one-out cross-validation in blue, and shuffled data in grey. *=p<0.001 vs. shuffled data.

Finally, we used SVM-based pattern classification to examine if we could predict dementia in PD from these first 3 principal components. Critically, all classification was binary and performed within PD patients. We found that SVMs could accurately distinguish PDD (16 patients) from PD patients (40 patients; AUC=0.75; Fig. 5C). This was significantly higher than AUCs in shuffled data (shuffled AUC =0.50; p<0.001; Fig. 5G) and higher than AUCs for detection of PDMCI from PD (AUC=0.65) and from PD (AUC=0.49). These findings provide insight into the nature of midfrontal delta/theta rhythms and into cognition in PD. Taken together, our results suggest that evoked midfrontal activity can identify cognitive dysfunction within PD patients.

## Discussion

We tested the hypothesis that diminished midfrontal delta/theta rhythms are associated with cognitive dysfunction in PD. Across three cognitive tasks in 100 PD patients, common features of midfrontal delta rhythms were decreased with PD-related cognitive dysfunction. Shared midfrontal variance was succinctly captured by data-reduction via PCA, which was used in turn to predict phenotypic expression on the MOCA or tests of executive function. These results are particularly remarkable when considering they relied only upon a single midfrontal scalp EEG electrode at Cz. Taken together, our results indicate that cue-evoked midfrontal delta/theta rhythms are highly predictive of cognitive dysfunction in PD.

Our work is in line with prior studies from our group and others documenting reliable PD-related decreases in midfrontal delta/theta rhythms.^15,16,34,42,43,53^ Here we extend this work to demonstrate that PD patients with cognitive dysfunction have progressive deficits in these low-frequency cortical rhythms, with PDD patients having most attenuated midfrontal delta/theta activity. These deficits are not readily explained by motor deficits, as patients with low UPDRS values and better motor function had higher PC1 scores, and this relationship was distinct from the relationship between PC1 and executive function. Our findings have mechanistic significance because midfrontal delta/theta activity signals need for cognitive control in response to novelty, errors, timing, or conflict across species.^13,21,42,54–58^

Low-frequency delta/theta bursts can powerfully engage cortical and subcortical brain networks,^12,19,20,59,60^ and synchronize single neurons in local and distant brain regions^18,21,61,62^ involved in the details of cognitive processing. Our work suggests that cognition is dysfunctional in PD patients because they fail to modulate cue-evoked midfrontal delta/theta rhythms to engage in cognitive operations, resulting in cognitive dysfunction and ultimately contributing to dementia. Supporting this idea is evidence that prefrontal ∼4 Hz stimulation can boost cognitive performance in PD and in PD rodent models,^20,63–65^ although much work remains to develop this idea into viable therapy for PD patients.

Critically, our results also suggest that midfrontal delta/theta deficits in PD patients are not strongly related to cognitive control, as noted by unreliable Group effects for both response times and EEG metrics on congruent vs. incongruent trials or on oddball vs. standard trials. The finding of maintained control-specific response time slowing is in line with prior work,^18,43^ and suggests that low-frequency cortical deficits may reflect orienting or engagement signals rather than specific control-related processes *per se*.^34,66^ Because cognitive dysfunction in PD involves specific mechanistic themes of dopamine dysfunction, cholinergic dysfunction, and associated cortical dysfunction,^67,68^ these findings work provide insight into the nature of these midfrontal rhythms.

In addition, our results advance the idea of midfrontal delta/theta activity as a candidate biomarker for cognitive deficits in PD. Midfrontal theta is well understood as a candidate mechanism for cognitive control with well-defined psychometric properties.^13,69,70^ The lack of strong control-related effects suggest that even a low-salience imperative cue requiring a response might trigger midfrontal delta/theta activity and be attenuated in PD.^53^ We note that our tasks are simple and time-efficient, and can performed by PD patients with marked cognitive dysfunction as well as in the operating room.^18,20^ Fascinatingly, response-time effects are primarily observed in PDMCI/PDD, which may underlie classic behavioral studies in PD that simply describe behavioral slowing.^71,72^ Our behavioral data imply that PD patients with cognitive dysfunction have the most pronounced behavioral slowing. We find strong effects of PD-related cognitive dysfunction on both behavior and mid-frontal delta/theta activity, and some event-related potentials (i.e., P3a), making these tasks ideal for the development of future biomarkers.^16,44^ Pattern classification of this burst of EEG activity from a single scalp electrode separated PD patients from those with dementia, which could be of great utility in developing objective neurophysiological biomarkers, particularly when movement disorders and/or neuropsychological expertise is in short supply.

Our study is limited by the number of tasks and assays we performed, although we were constrained by patient-related fatigue, particularly in groups with PDMCI/PDD. Furthermore, EEG is inherently limited in its spatial resolution, although the temporal resolution permits detailed time-frequency analysis. Further, the ubiquity and availability greatly enhance the usefulness of EEG and could make these midfrontal signals compatible with closed-loop control for future brain stimulation or neuromodulatory therapies to improve cognitive deficits in PD.

In summary, our data demonstrate that midfrontal delta signals were reliably attenuated in PD-related cognition, and could be used to predict cognitive dysfunction in PD. These findings imply that PD-patients have cognitive deficits because they fail to engage basic orienting processes. This mechanistic insight could inspire future work on fundamental processes that fail in PD patients, and could inspire new biomarkers as well as novel rehabilitation and neuromodulation strategies targeting cognition in PD.

## Data Availability

All data produced in the present study are available upon reasonable request to the authors.

## Funding

This data was supported by NIH P20NS123151 and R01NS100849 to NSN, NRSA F32 AG069445-01 to RC.

## Competing interests

The authors report no competing interests.

## Supplementary material

Supplementary material is available at *Brain* online

## Supplementary Materials

**Figure S1:**
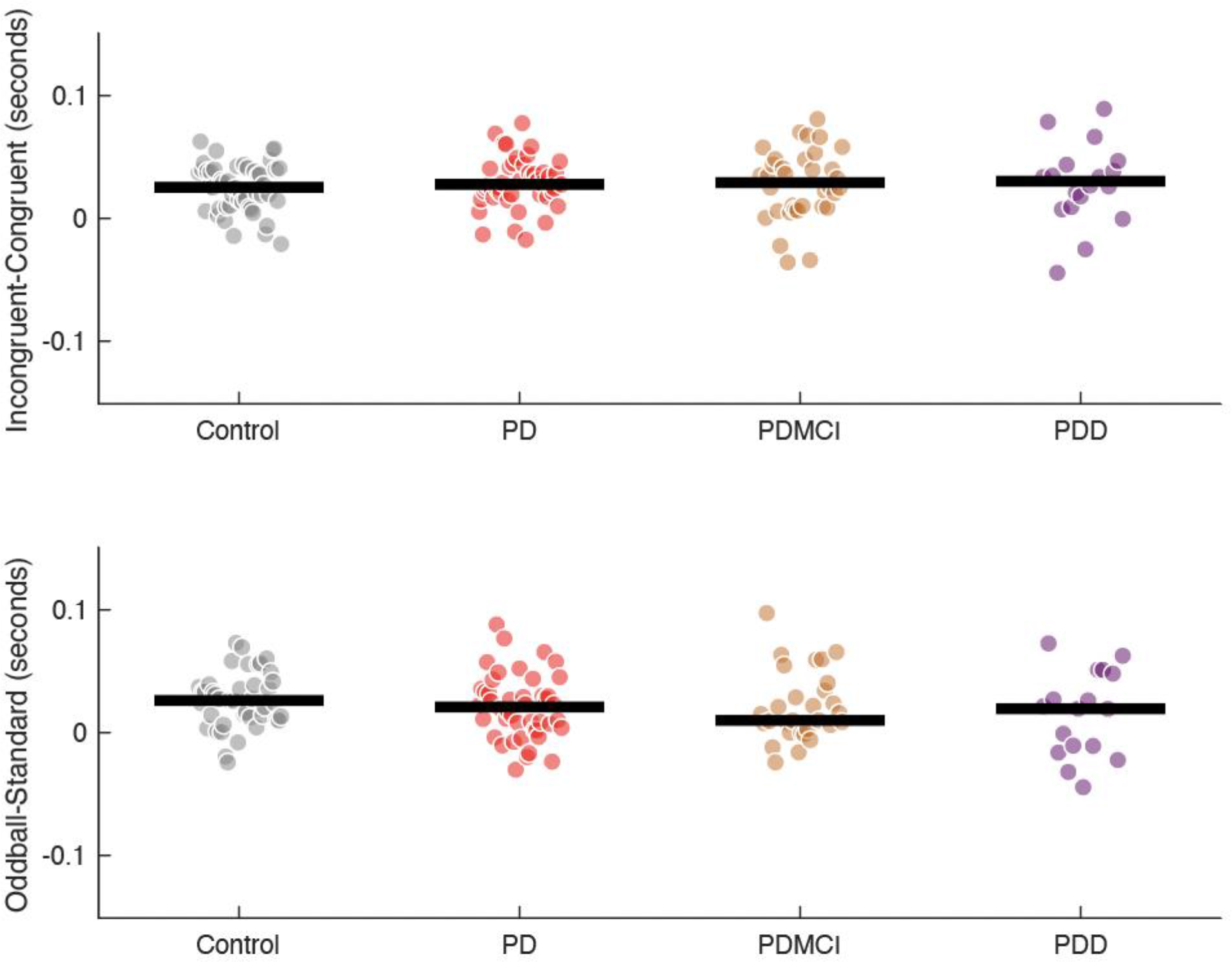
Control-related slowing on Simon and oddball tasks. Top: differences in response latencies (in seconds) on incongruent vs. congruent trials for control (gray), PD patients (red), PDMCI (orange), and PDD (plum) from the Simon task. Bottom: differences in response latencies (in seconds) on oddball vs. standard trials for control (gray), PD patients (red), PDMCI (orange), and PDD (plum) from the oddball task.

**Figure S2:**
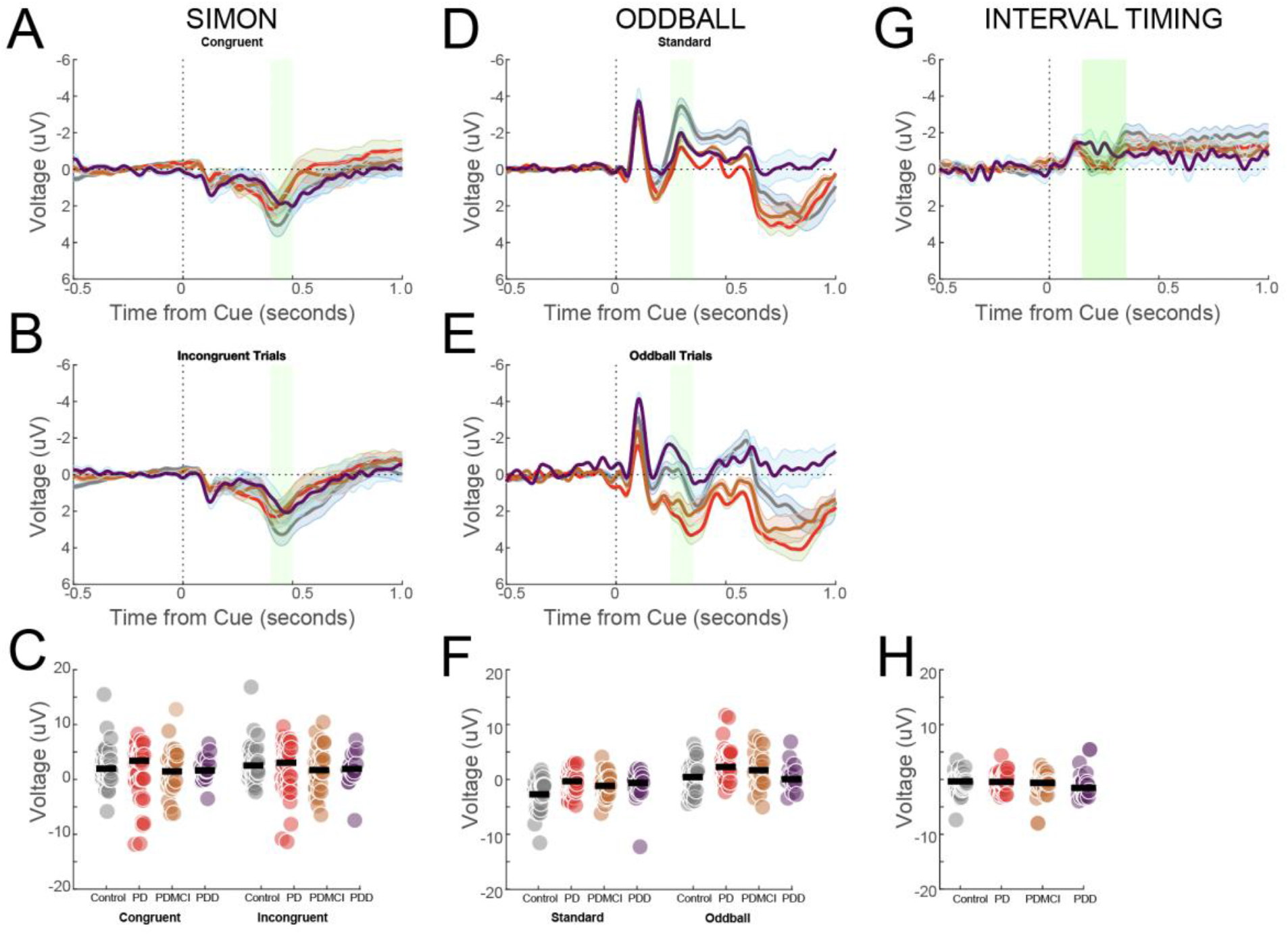
Event-related potential components are not reliably affected by cognitive dysfunction in PD: ERP voltage during the Simon task from electrode Cz for A) congruent and B) incongruent trials for response times for congruent and incongruent trials for 48 control (gray), 47 PD (red), 34 PDMCI (orange), and 18 PDD (plum) patients; the black bar is the median. C) Average amplitudes across patients; the black bar is the median. ERP voltage during the oddball task from electrode Cz for D) standard and E) oddball trials for 41 control, 43 PD, 29 PDMCI, and 18 PDD patients. F) Average amplitudes across patients; the black bar is the median; *main effect of trial type; # main effect of group. G) ERP voltage during the Interval Timing task from electrode Cz for 43 control, 43 PD, 32 PDMCI, and 19 PDD patients. H) Average amplitudes across patients; the black bar is the median. Note that ERPs are plotted with negative voltages up.

**Figure S3:**
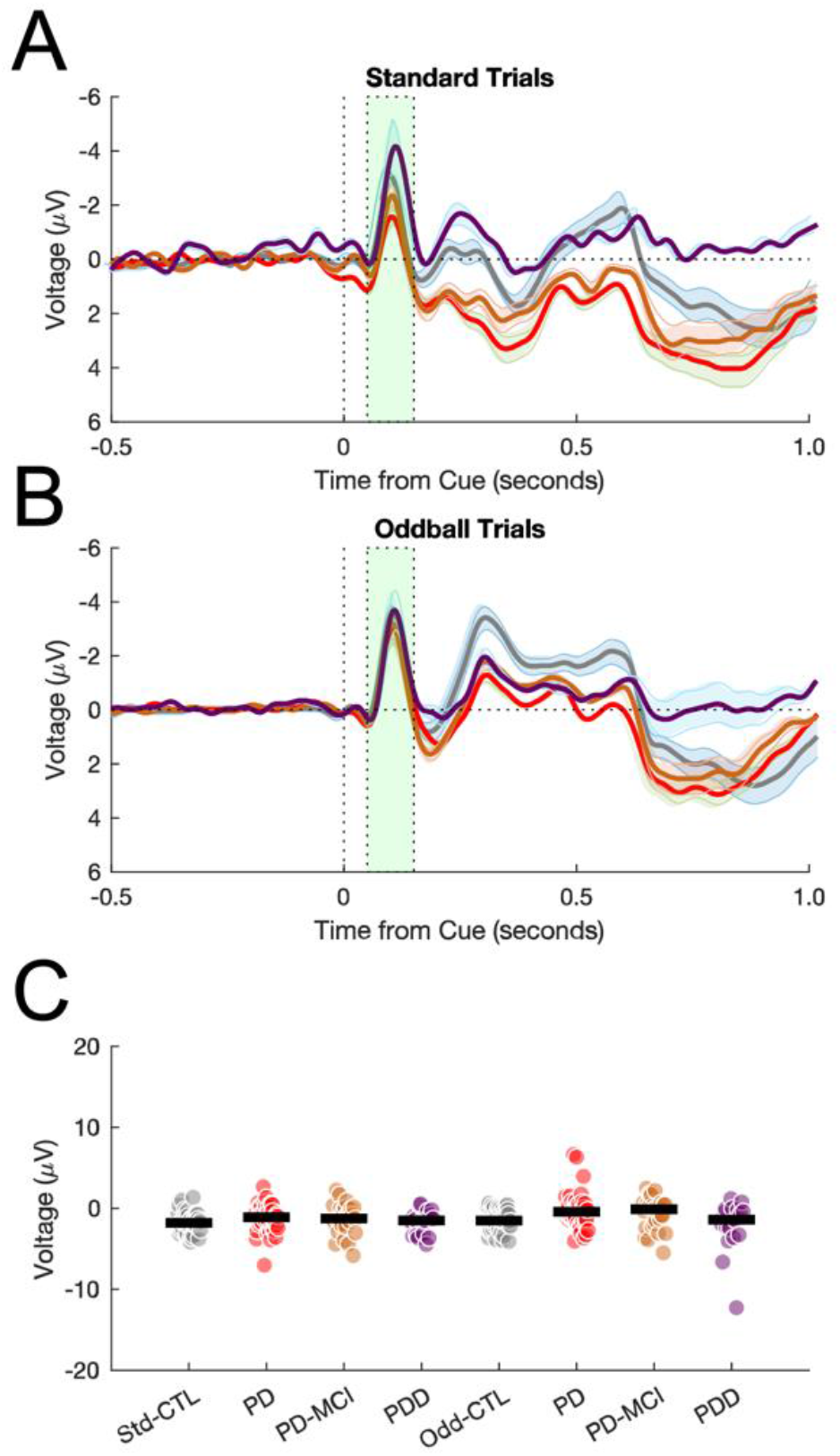
Oddball ERPs. A) ERP voltage during the oddball task from electrode Cz for standard and B) oddball trials C) Average amplitudes from 0.05 – 0.15 seconds across patients; the black bar is the median for 41 control, 43 PD, 29 PDMCI, and 18 PDD patients.

**Figure S4:**
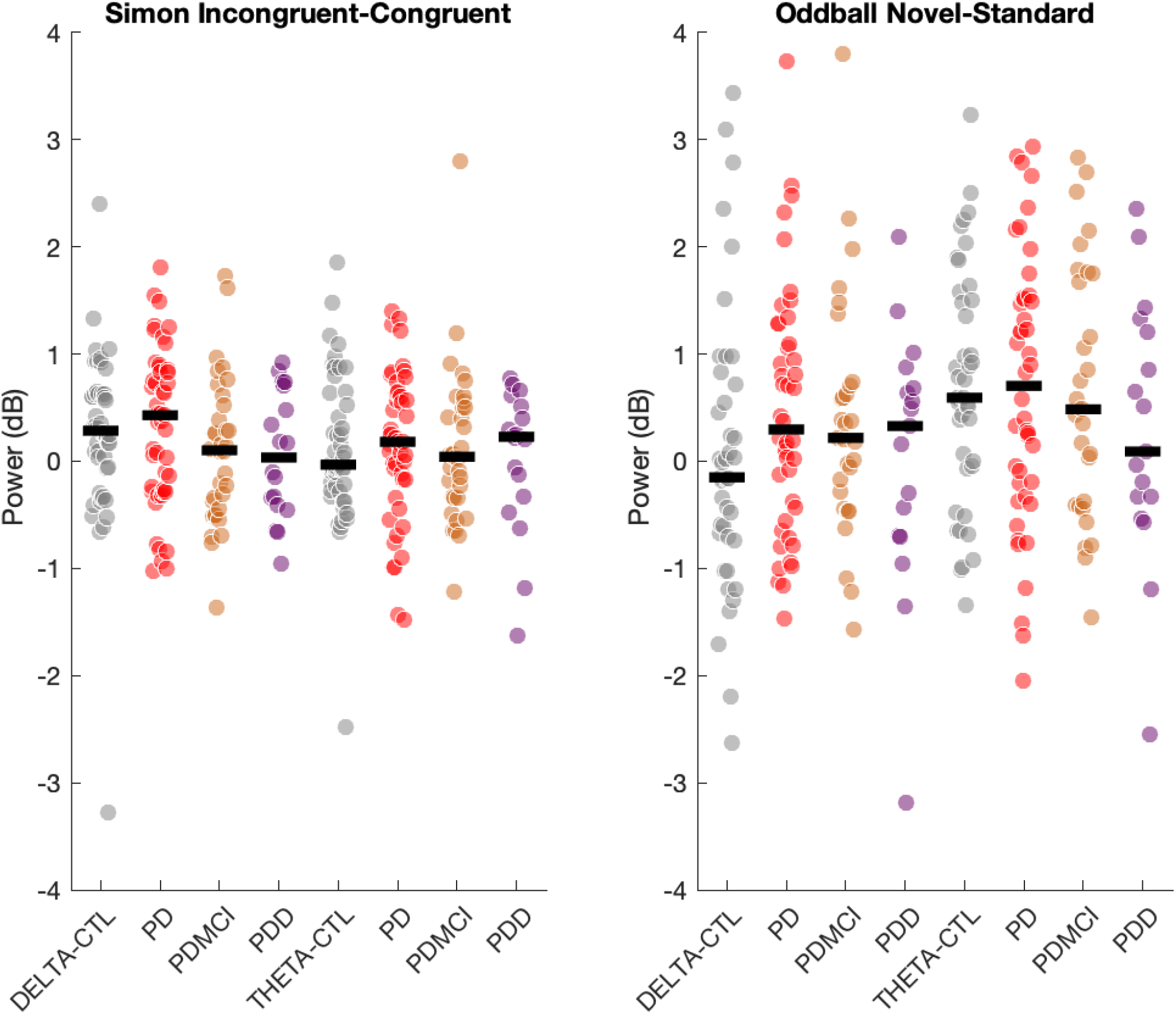
Control-related power. Power from electrode Cz corresponding to ROIs in Figure 3 during the Simon task on incongruent - congruent trials (left), and during the oddball task on oddball – standard trials (right).

**Figure S5:**
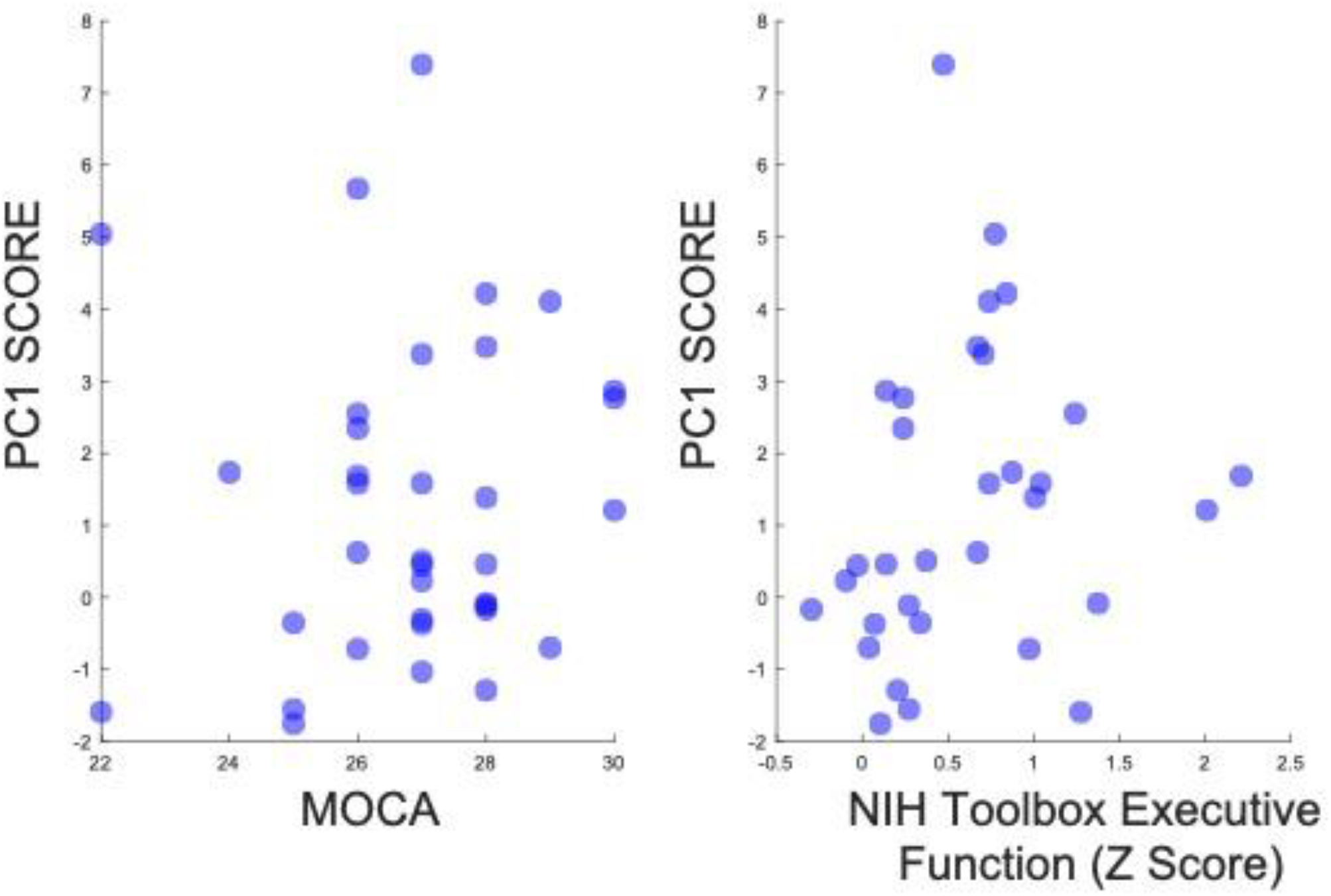
PC1 correlations with MOCA and NIH Toolbox Executive Function for 35 control participants. Neither relationship was statistically significant.

## Notes

### Competing Interest Statement

The authors have declared no competing interest.

### Author Declarations

All research protocols were approved by the University of Iowa Human Subjects Review Board (IRB# 201707828).

